# The science of child and adolescent mental health in Mozambique: a nationwide systematic review

**DOI:** 10.1101/2025.09.14.25335725

**Authors:** Helena Mutede Cutótua Daniel, Lauro Estivalete Marchionatti, Jessica Azevedo Veronesi, André Cardoso Campello, Luis Augusto Rohde, Caio Borba Casella, Peter Raucci, Afonso Mazine Tiago Fumo, Jair Mari, Giovanni Abrahão Salum, Lidia Gouveia, Zeina Mneimneh

## Abstract

**Introduction:** Mozambique is home to 18 million children and adolescents largely underrepresented in mental health research. We conducted a systematic review of evidence-based resources addressing this population.

**Methods:** We included prevalence estimates, assessment instruments, and interventions on mental health-related outcomes in Mozambican samples aged 0-19. We searched PubMed, Web of Science, PsycINFO, CINAHL, Google Scholar, African Index Medicus, and local catalogues. Extraction followed COSMIN and Cochrane manuals (PROSPERO: CRD420251016208).

**Results:** The search yielded 58 prevalence studies, 26 reports on 35 instruments, and 7 trials. Most research focused on psychosocial adversities and neurodevelopment in vulnerable populations. The prevalence of mental disorders were estimated in two surveys in Nampula (ADHD: 13.4%; N=748, 6-18 years) and Maputo (anxiety disorders: 17.5%; major depression episode: 8.5%; disruptive behavior disorder: 6.6%; ADHD: 3.3%; N=486, 12-19 years). Nationally-representative adolescent surveys report health determinants (physical/emotional violence anually: 24.86%; girls suffering intimate partner violence: 21.11%; lifetime sexual abuse: 16.55%). National suicide rates were highest among 15-19-year-olds (1.53 per 100,000). Only three instruments were culturally-adapted with reliable psychometrics, enabling screening of anxiety and general symptoms. Unadapted tools performed poorly. One randomized trial reported a cost-effective intervention for neurodevelopment among high-poverty preschoolers.

**Discussion:** Children and adolescents in Mozambique face significant psychosocial adversity, with a high estimated burden of mental disorders. Prevalence data remains limited to two localized samples. There are few tools evaluating mental conditions, and culturally-sensitive approaches are warranted. It is essential to strengthen local academic capacity.

## Introduction

The emerging field of global mental health requires strengthening resources in lower-middle income countries (LMIC), where approximately 90% of the world’s population under 19 lives but remain underrepresented in research data.^1^ A recent global analysis shows the mental health data gap is especially pronounced across sub-Saharan African countries, with a near absence of nationally representative data on children and adolescents.^2^ Mozambique, located in Southern Africa, is an illustrative case. The country is home to more than 18 million of children and adolescents, representing over 55% of the total demographic composition.^1^ It is estimated that the majority of these youth live below the poverty line, with socioeconomic vulnerabilities extending to climate-related disasters, exposure to violence and conflict, and the HIV/AIDS epidemic.^3^ Their mental health and psychosocial development have gained increased attention from the national government and international agencies, with the Ministry of Health including children and adolescents as a priority in the Strategy and Action Plan for Mental Health 2016–2026.^4^

Mental health resources in Mozambique are critically strained. The country has fewer than 500 specialists (including psychiatrists, psychologists, and psychiatric technicians), with the first child and adolescent psychiatrist being trained in 2021.^5,6^ This shortage is reflected in the limited volume of research produced locally. A 2019 systematic review identified just 130 mental health–related publications from Mozambique, revealing substantial gaps in epidemiological data.^7^ Despite these constraints, research output appears to be increasing: a PubMed search using the terms “mental health” AND “Mozambique” yields 189 records, 144 of which have been published since 2020. Some of this work explicitly focuses on bridging gaps in evidence-based resources for children and adolescents, including global and national surveys and work validating assessment instruments.^8^

In non-Western contexts, synthesizing and appraising the research base is essential to enhance the reach and availability of mental health resources. National-level compilations on child and adolescent mental health have been conducted in countries such as Greece,^9^ Brazil,^10^ and South Africa,^11^ providing valuable resources to inform practice, policy, and research. This study aims to synthesize the scientific literature on child and adolescent mental health in Mozambique. We conducted a systematic review of prevalence surveys, assessment instruments, and interventions addressing mental health–related outcomes in this population. Our goal is to present a comprehensive overview of evidence-based resources available in the country and to take practical insights for public health.

## Methods

We draw on national reviews conducted in Greece, Brazil, and South Africa, with a comprehensive search strategy followed by classification of studies into three independent arms (prevalence, assessment instruments, and interventions).^9–11^ The protocol has been published in PROSPERO [CRD420251016208]. All research conducted in Mozambique requires registration, and we obtained approval from the Ethics Committee of the Faculdade de Medicina da Universidade Eduardo Mondlane (UEM) and the Hospital Central de Maputo [CIBSFM&HCM/42/2025]. This review follows the Preferred Reporting Items for Systematic Reviews and Meta-Analyses (PRISMA) guidelines and the PRISMA-COSMIN manual for reporting systematic reviews of measurement instruments.^12,13^

### Inclusion criteria

Our scope was the mental health of children and/or adolescents in Mozambique considering outcomes in a broad sense (e..g: mental disorders, substance use, suicidality, life adversities, childhood maltreatment and neglect, bullying, quality of life, wellness, neurodevelopment, learning difficulties, literacy, stigma). For eligibility, studies had to report specific data for samples in Mozambique with an average age up to 19-years-old (regardless of outliers).

Eligible studies were classified into one or more of three review arms based on the following criteria: (1) prevalence studies included community, school or clinical surveys using structured instruments, questionnaires, or epidemiological registries; (2) assessment instruments included studies that developed, translated or validated tools, further including studies that solely applied an instrument if that was the unique report of the tool in the country; (3) interventions studies with experimental (or quasi-experimental) designs, as well as translation, adaptation, or development of intervention programs. We included peer-reviewed articles, academic letters, book chapters, theses and dissertations, and institutional reports, without restriction of language or date. For repeated datasets, we included the most recent and/or most comprehensive report. We excluded multi-country studies that did not provide specific data for Mozambique, as well as general population studies without specific data for children or adolescents. Conference abstracts were not eligible.

### Search strategy and screening

We searched multiple data sources from inception to December 4th 2024: (1) international databases, including PubMed, Web of Science, CINAHL, PsycInfo, and Google Scholar; (2) the African Index Medicus; (3) local university repositories from Universidade Lúrio (UniLúrio) and Universidade Eduardo Mondlane (UEM); (4) publications from the National Health Institute (Instituto Nacional de Saúde – INS); and (5) the local health journals, Revista Moçambicana de Ciência de Saúde (RMCS) and Revista Médica de Moçambique (RMM).

A comprehensive query combining terms related to children/adolescents, mental health, and Mozambique was adapted to the syntax and language (English and/or Portuguese) of each database (see Supplementary Table 1). First, records were retrieved from PubMed, Web of Science, CINAHL, and PsycInfo. Following deduplication, two independent authors conducted primary screening (based on titles and abstracts) and a single reviewer performed full-text screening. Second, we searched Google Scholar as a complementary source, adapting the query to its crawler-based engine using simplified terms in English and Portuguese. A single author screened results until 100 consecutive entries produced no new relevant studies. Next, regional and local data sources were also screened by a single author. African Medicus Index was searched for records containing “Mozambique” or “Moçambique”; the UEM repository was searched with Portuguese terms related to children and adolescents; and the full catalogues of the remaining sources (INS, UniLúrio, RMCS, RMM) were manually verified.

The research team conducted a pilot calibration on 10% of the dataset. Regular meetings were held to ensure consistency and shared understanding across reviewers throughout screening and extraction. Single-reviewer screening procedures had an emphasis on sensitivity over specificity to maximize inclusion, and 20% of the records were randomly verified by a second reviewer (LEM). This author also resolved discrepancies arising from two author independent screening.

The review was managed using Rayyan software. Potential duplicates were detected, automatically removing pairs with an overall similarity index above 95% or a 100% match in title or DOI. Remaining candidates were manually verified.

### Data extraction

Data collection was conducted independently for each review arm, following specific methodological guidelines. A single reviewer performed the initial extraction, and all datasheets were subsequently verified by LEM.

Prevalence studies followed procedures established in a widely cited systematic review on child and adolescent mental health disorders.^14^ Risk of bias is assessed using a validated tool that evaluates external and internal validity, analytical bias, and population representativeness.^15^ Supplementary Table 2 presents extraction fields, with each estimate from each study representing a separate data entry.

For assessment instruments, we followed the Consensus-based Standards for the Selection of Health Measurement Instruments (COSMIN) for data extraction, quality assessment, and psychometric appraisal.^16^ Supplementary Table 3 shows extraction fields, collecting information for each instrument reported in a study on a separate line. Next, data is classified using psychometric criteria outlined in Supplementary Table 4. Finally, we generate a summary for each instrument by aggregating data across studies (see codes in Supplementary Table 5).

Intervention studies were extracted following the Cochrane Manual for Systematic Reviews of Interventions.^17^ Methodological quality was assessed with the revised Cochrane tool for randomized trials (RoB 2) and the Joanna Briggs Institute (JBI) checklist for non-randomized designs.^18,19^ Extraction fields are detailed in Supplementary Table 6.

## Results

Figure 1 presents a flowchart with screening outcomes, and Supplementary Table 7 lists reasons for exclusion. After screening over 4,435 records, there were 76 unique publications included across three areas: 58 prevalence reports on 213 estimates, 26 studies on 35 assessment instruments, and 8 studies describing 7 intervention trials. The full dataset is available in Supplementary File A. Prevalence data is summarized in Table 1 and Table 2. Assessment instruments are shown in Table 4 and Supplementary Table 8. Intervention trials are reported in Table 4.

**Figure 1.**
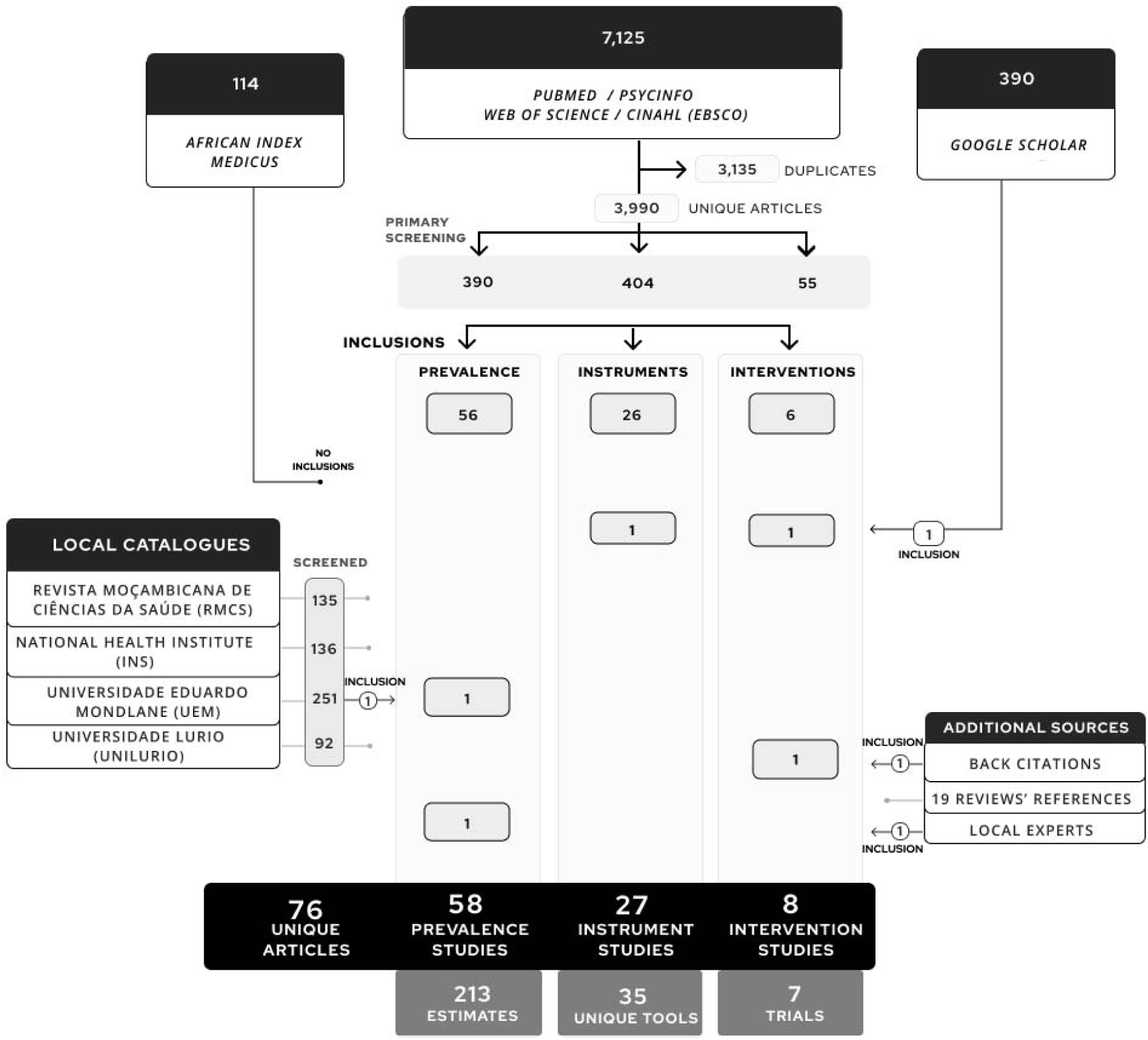
Flowchart of inclusions

**Table 1.** Prevalence studies with structured clinical assessment or clinical registers.

| CLINICAL ASSESSMENT WITH STRUCTURED CRITERIA FOR DIAGNOSIS |  |  |  |  |  |
| --- | --- | --- | --- | --- | --- |
| Study | Methods | Sample (N)<br>Description | Location<br>Year | Diagnostic criteria | Findings |
| 62,63 | Cross-sectional<br>School-based<br>Probabilistic | N = 748<br>6 to 18 years-old<br>54.6% female | Nampula<br>2022 - 2023 | Kiddie Schedule for Affective Disorders and Schizophrenia for School-aged Children<br>DSM-5 criteria<br>Interviews were applied to patients who screened positive with SNAP-IV rated by teachers and caregivers | Current prevalence<br>ADHD: 13.4% (95% CI 11.5-19.2)<br>SNAP-IV positive screening: SNAP-IV teacher or caregiver: 10,16% |
| 8,64 | Cross-sectional<br>School-based<br>Probabilistic | N = 486<br>12 - 19 years-old<br>66.2% female | Maputo<br>2019 | Miniature International Neuropsychiatric Interview for Children and Adolescents (MINI-KID)<br>DSM-4 and ICD-10 criteria | Current prevalence:<br>Anxiety disorders<sup>1</sup> 17.5%<br>Major Depressive Disorder Episode: 8.5%<br>Disruptive Behavior Disorder<sup>2</sup>: 6.6%<br>ADHD: 3.3%<br>Suicidal thoughts (past month): 16,42%<br>Suicidal behavior (past month): 6,16%<br>Suicide attempt (lifetime): 8,89% |
| 65 | Case-control on a cohort of survivors of invasive Group B Streptococcus infant<br>Controls identified via Health and Demographic Surveillance System | N = 110<br>1 - 20 years-old<br>54.5% female<br>Control group matched for survivors of invasive bacterial infection during early infancy | Maputo<br>2019 - 2021 | Battery of neurodevelopmental tests:<br>Malawi Developmental Assessment Tool (MDAT)<br>Cambridge Neuropsychological Test Automated Battery (CANTAT)<br>WHO and Global Burden of Disease (GBD) criteria for neurodevelopmental impairment | Neurodevelopmental impairment (moderate to severe): 0.9%<br>Neurodevelopmental impairment (mild): 18.2%<br>Cognitive impairment (moderate to severe): 4,8%<br>Cognitive impairment (mild): 14.4%<br>*Obs: No clinical cases were identified through the Child Behavior Checklist (CBCL), which was attributed to a poor* |

|  |  |  |  |  | *cultural fit of the instrument* |
| --- | --- | --- | --- | --- | --- |
| **REGISTER-BASED STUDIES** |  |  |  |  |  |
| Study | Methods | Sample representativeness | Location<br>Year | Diagnostic criteria | Findings |
| <sup>23</sup> | Record book of autopsies at Forensic Services | National population<br>All suicides should undergo autopsy and data was collected at all forensic central services attending Mozambique at the time | Central Hospitals of Maputo, Beira and Nampula<br>2014 - 2019 | Autopsy registers | Absolute number of suicides<br>• 5 - 19 years-old: 70<br>• 5 - 9 years-old: 2<br>• 10 - 14 years-old: 21<br>• 15 - 19 years-old: 47<br>*Suicide rates per 100,000:<sup>2</sup>*<br>• 5 - 19 years-old: 0.65<br>• 5 - 9 years-old: 0.05<br>• 10 - 14 years-old: 0.59<br>• 15 - 19 years-old: 1.53 |
| <sup>66</sup> | Record book of autopsies at a regional forensic service<br>Emergency visits records at a regional hospital | Sofala and adjacent catchment area of Beira Central Hospital | Beira Central Hospital in Sofala<br>2013 - 2014 | Autopsy registers of suicide<br>Clinical registers of emergency visits | Absolute number of suicides in (up to 18 years-old): 19<br>Emergency visits due to suicidality (up to 18 years-old): 36 |
| <sup>67</sup> | Data is collected from statistical tables of the The State of the World's Children 2016, which are currently unavailable | National<br>Data is currently unavailable in the dataset | Unclear | Unclear | Child labor: 22%<br>Child marriage (married by 15): 14%<br>Child marriage (married by 18): 48%<br>Child neglect (children with inadequate supervision): 33% |
| <sup>68</sup> | Clinical registers from the four regional referral hospitals in Mozambique | Nationwide<br>Age: 0 - 14 years | Hospitals in Maputo, Beira, Quelimane and Nampula<br>2019 - 2020 | Clinical registers of paediatric patients admitted with acute injuries | Injury hospital admission due to interpersonal violence:<br>- 2019: 5 cases<br>- 2020: 4 cases |
| <sup>69</sup> | Clinical registers from a regional hospital | Maputo Central Hospital catchment area<br>Age: 0 - 14 years | Maputo Central Hospital<br>2019 | Clinical registers of paediatric patients admitted with acute injuries | Hospital visit due to physical violence: 64 cases<br>Hospital visit due to sexual violence: 79 cases |
| <sup>70</sup> | Clinical registers from a regional hospital | Hospital Central da Beira catchment area<br>Age: 0 - 14 years | Hospital Central da Beira<br>2017 - 2018 | Clinical registers of paediatric patients admitted with acute injury | Hospital visit due to violent injury: 143 |
| <sup>71</sup> | Clinical registers from a regional hospital | Hospital Central de Maputo catchment area<br>Age: 0 - 18 years | Hospital Central de Maputo<br>2015 - 2020 | Clinical registers from the Forensic Service | Child Sexual Abuse victims: 2610<br>93.7% girls<br>Assaults at the victims' or perpetrator's home: 54.8%<br>Perpetrator identified as an acquaintance: 72.9%<br>Concomitant physical violence: 54.1%<br>Perpetrator did not use condom: 74.9% |
| <sup>72</sup> | Clinical sample<br>HIV-infected infants aged <12 months at the time of hospital admission who had never initiated ART | N = 27<br>Up to 1 year-old<br>2019 | Hospital Central de Maputo (HCM) and Hospital Central de Beira (HCB). | Clinical charts classified according to established guidelines for developmental milestones | Delayed developmental milestones: 58,06% |

<sup>1</sup> Anxiety Disorders groups Generalized Anxiety Disorder, Separation Anxiety, Social Phobia, Panic Disorder, and Agoraphobia. <sup>2</sup>Disruptive Behavior Disorder groups Conduct and Oppositional Defiant Disorders
<sup>2</sup> We estimated relative rates considering the median of Mozambique population in each age group, from 2014 to 2019, as per the United States Census Bureau:<sup>1</sup> *1.08 million (5-19 years); 4,2 million (5 - 9 years); 3,5 million (10 - 14 years); 3 million (15 - 19 years).*

**Table 2.**
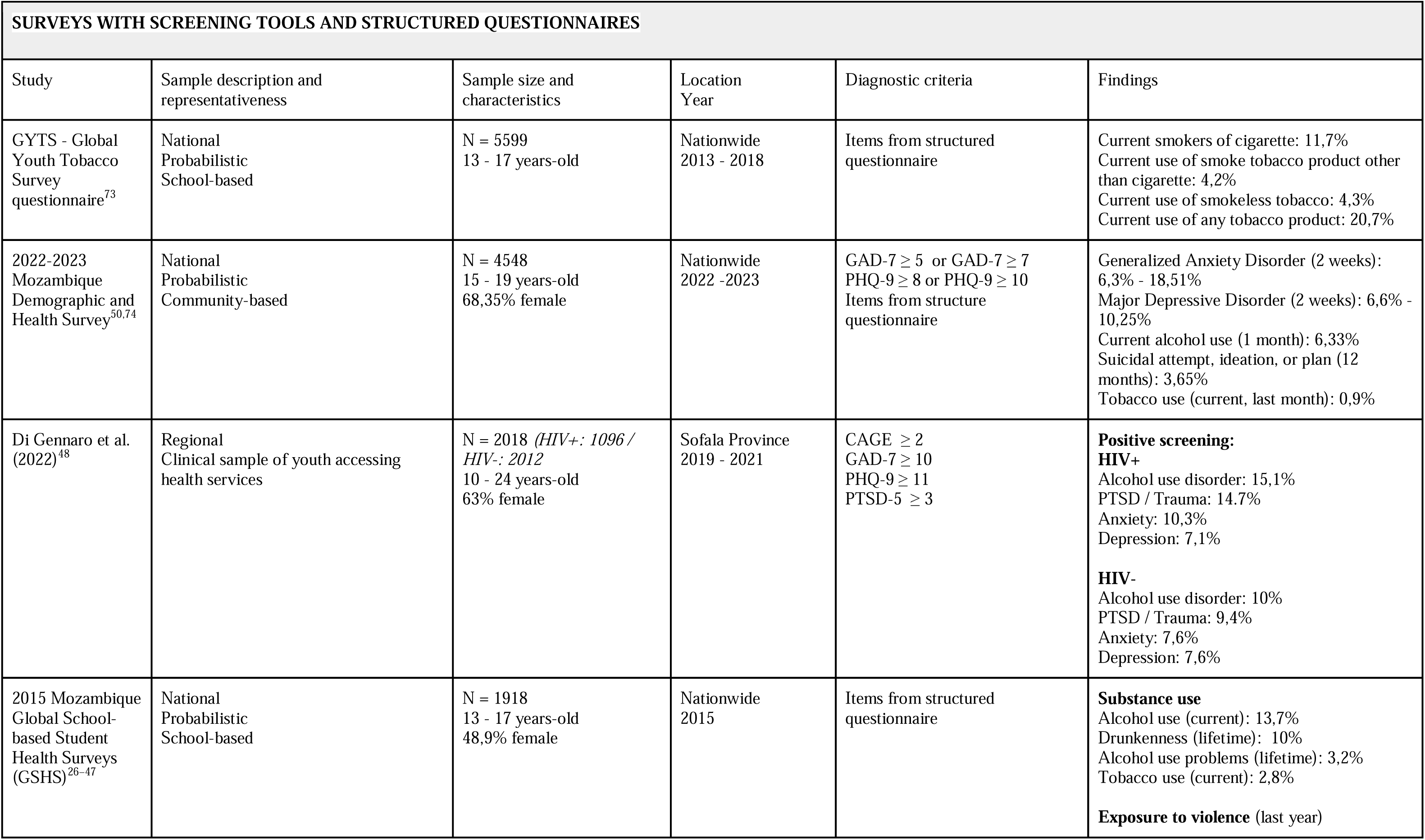
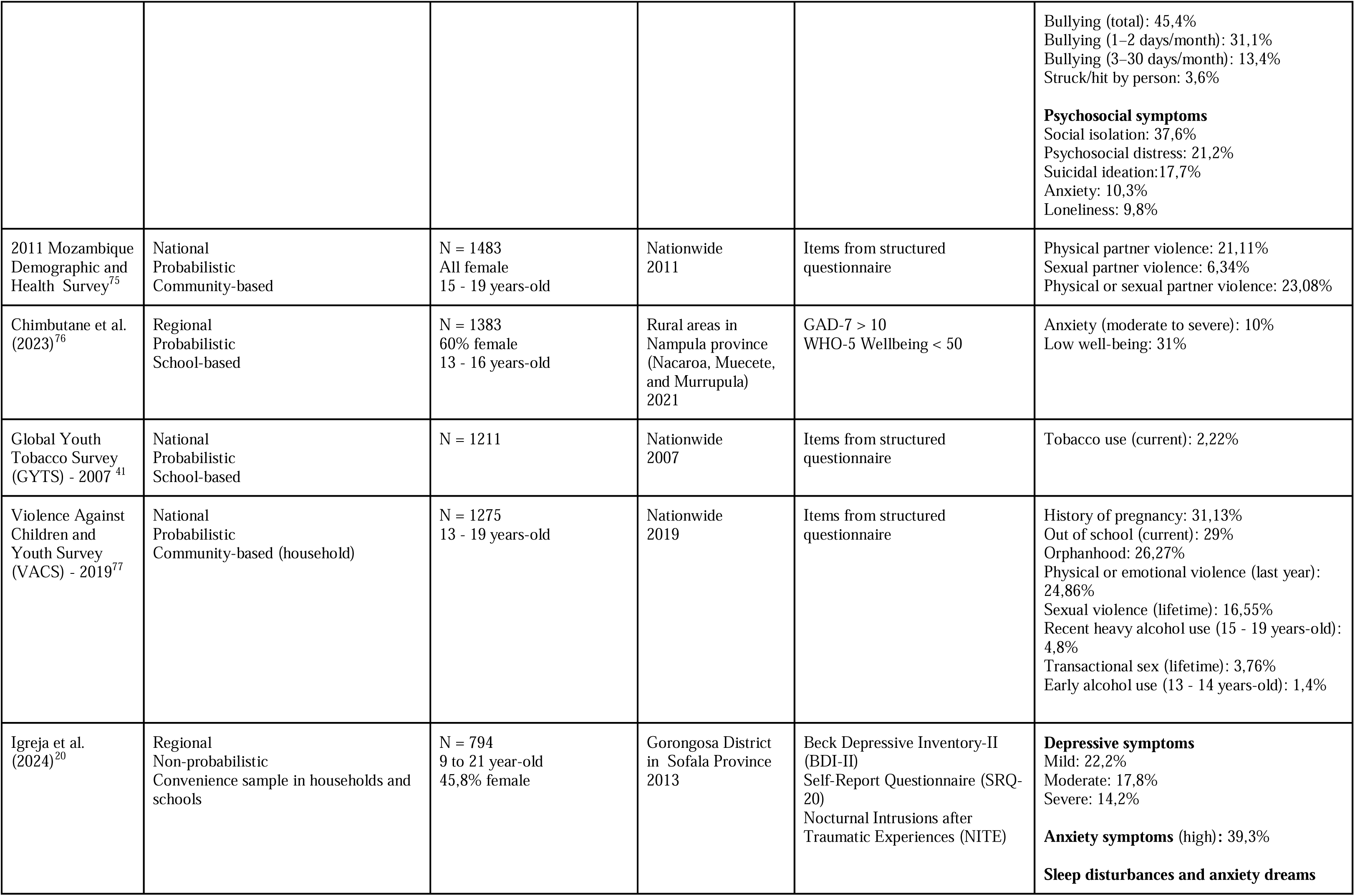
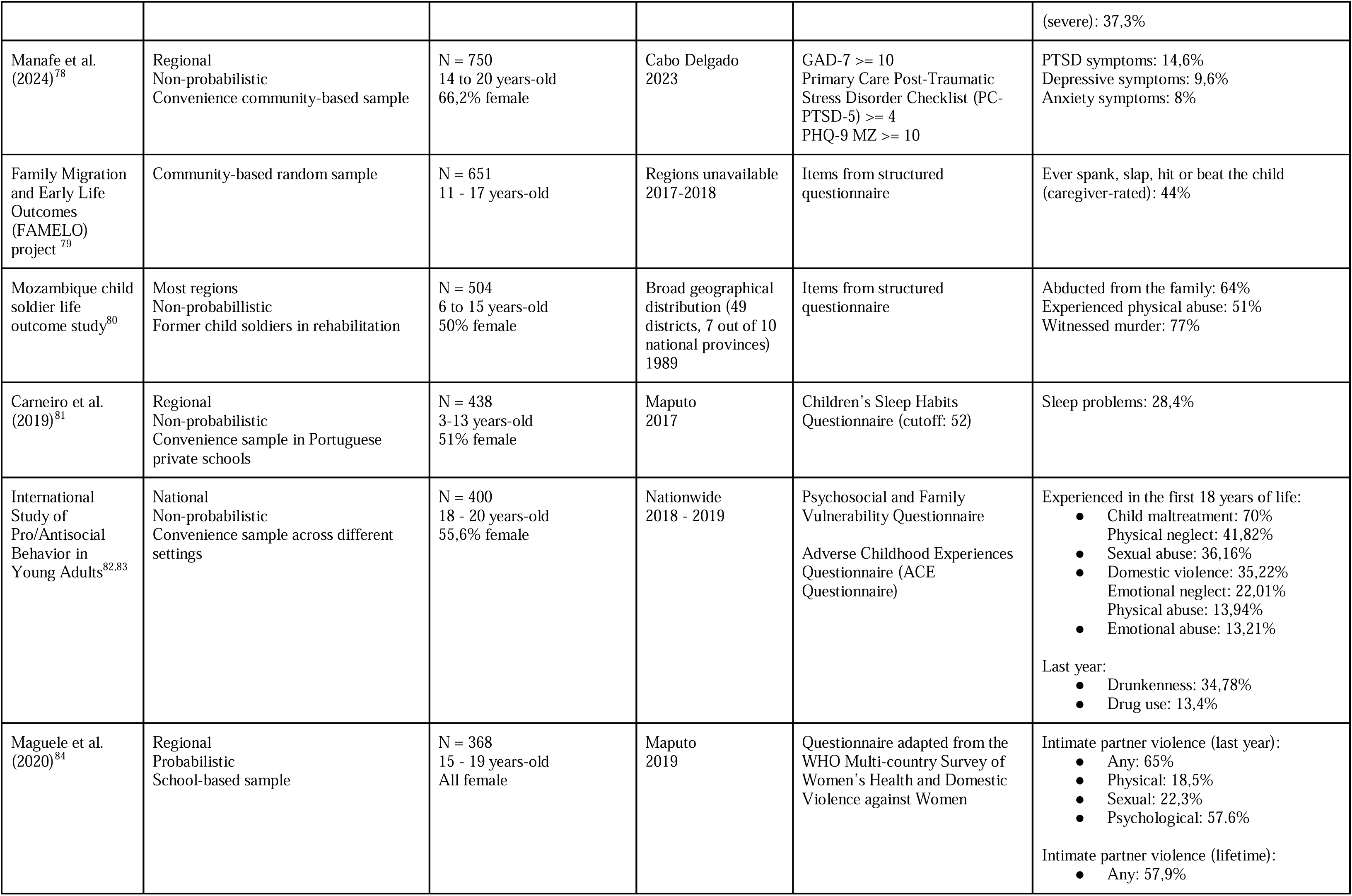
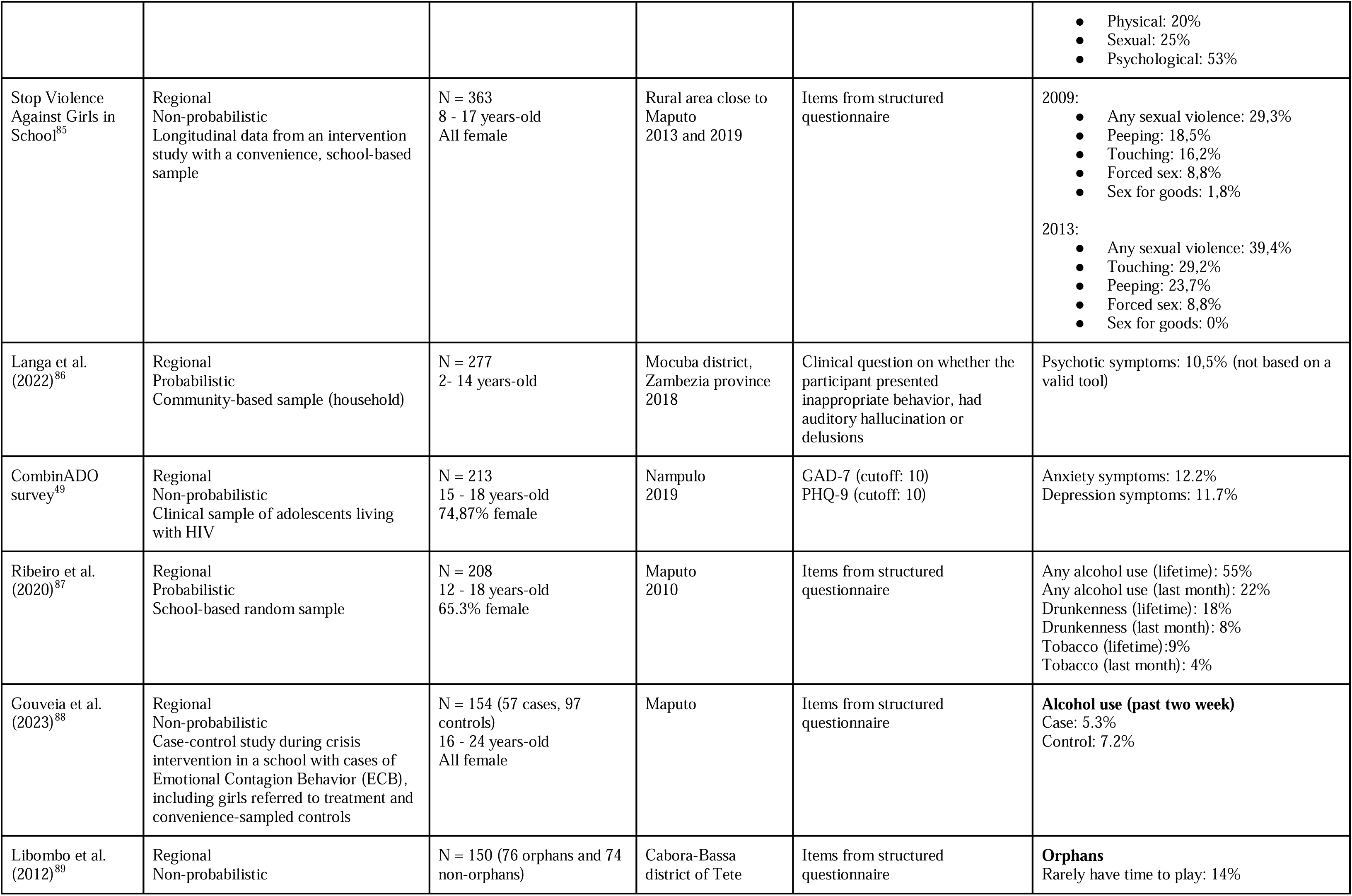
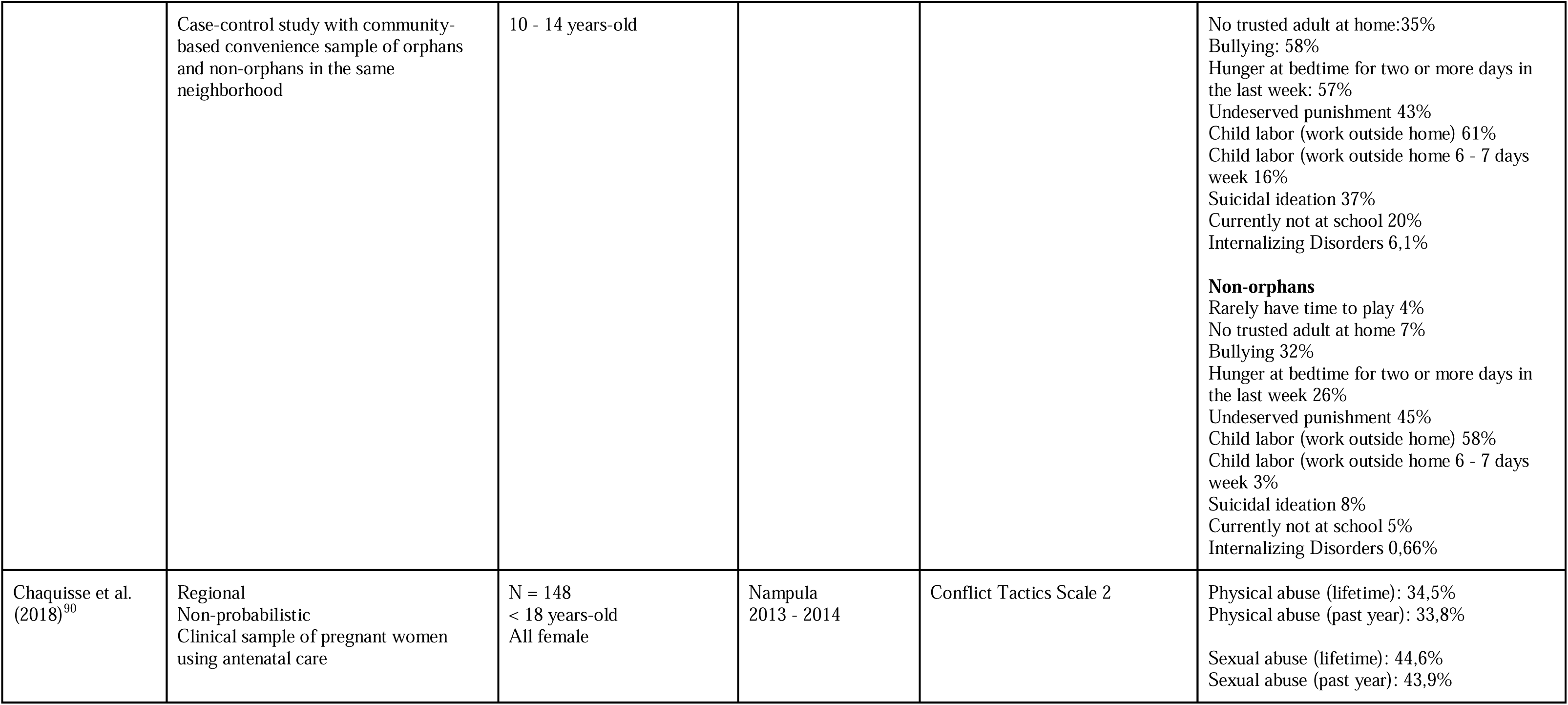
Prevalence studies with screening tools, surveys with questionnaires, and other methodologies.

### Overview of scientific production

Figure 2 presents the extent of evidence-based resources, while Figure 3 shows temporal trends of publications. There were 71 peer-reviewed articles, 3 theses/dissertations, and 2 institutional research reports. International authorship is predominant.^20^ Among the 29 articles with a Mozambican affiliation, just 18 credited a local institution in first authorship and only 6 in senior authorship. Two pHD theses were associated with international universities (Arizona State University and Universidade do Porto),^21,22^ with a single master dissertation produced in a local postgraduate program (UEM).^23^ Institutional research reports were published by World Bank^24^ and ActionAid.^25^

**Figure 2.**
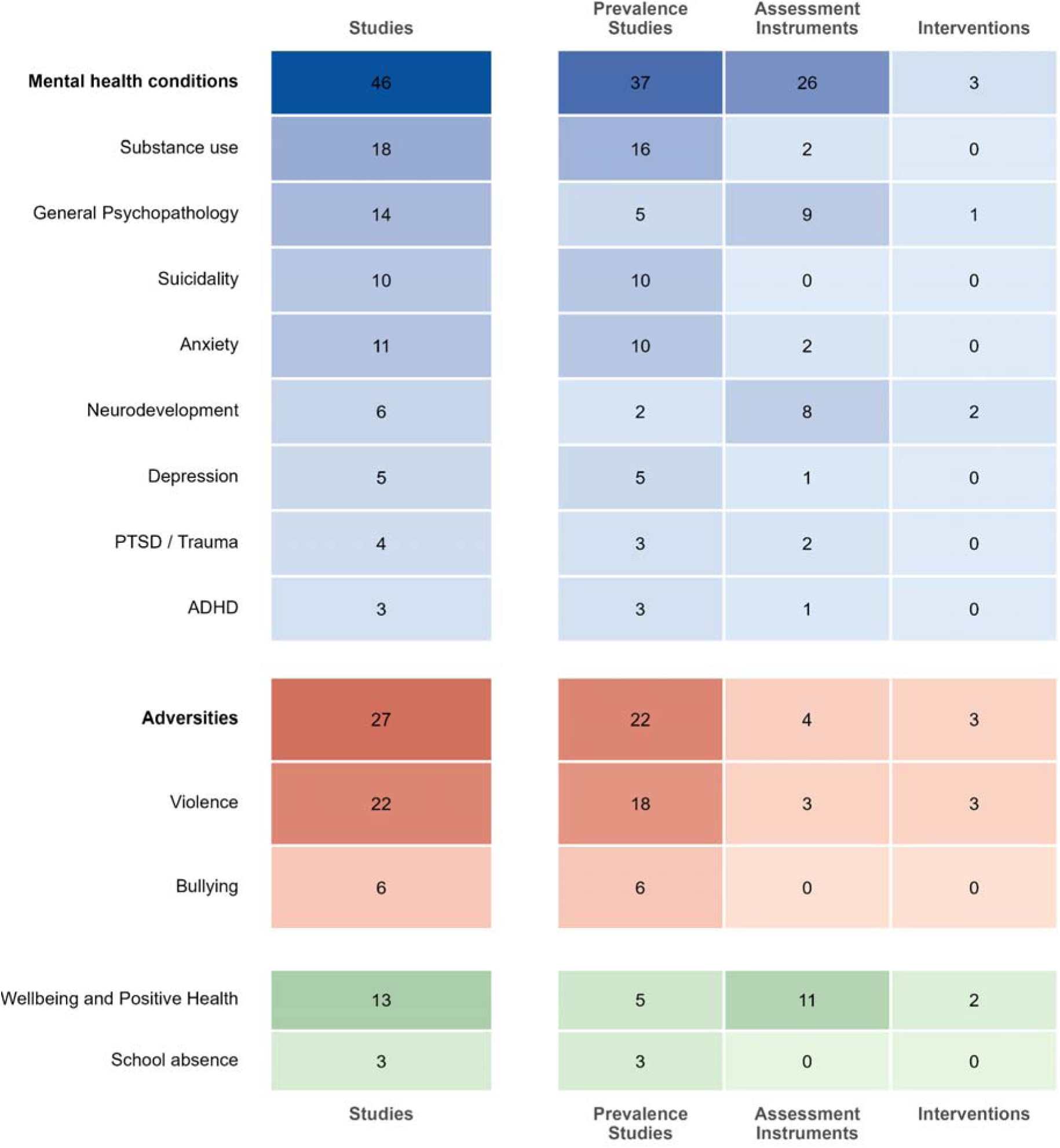
Availability of evidence-based resources per diagnostic category

**Figure 3.**
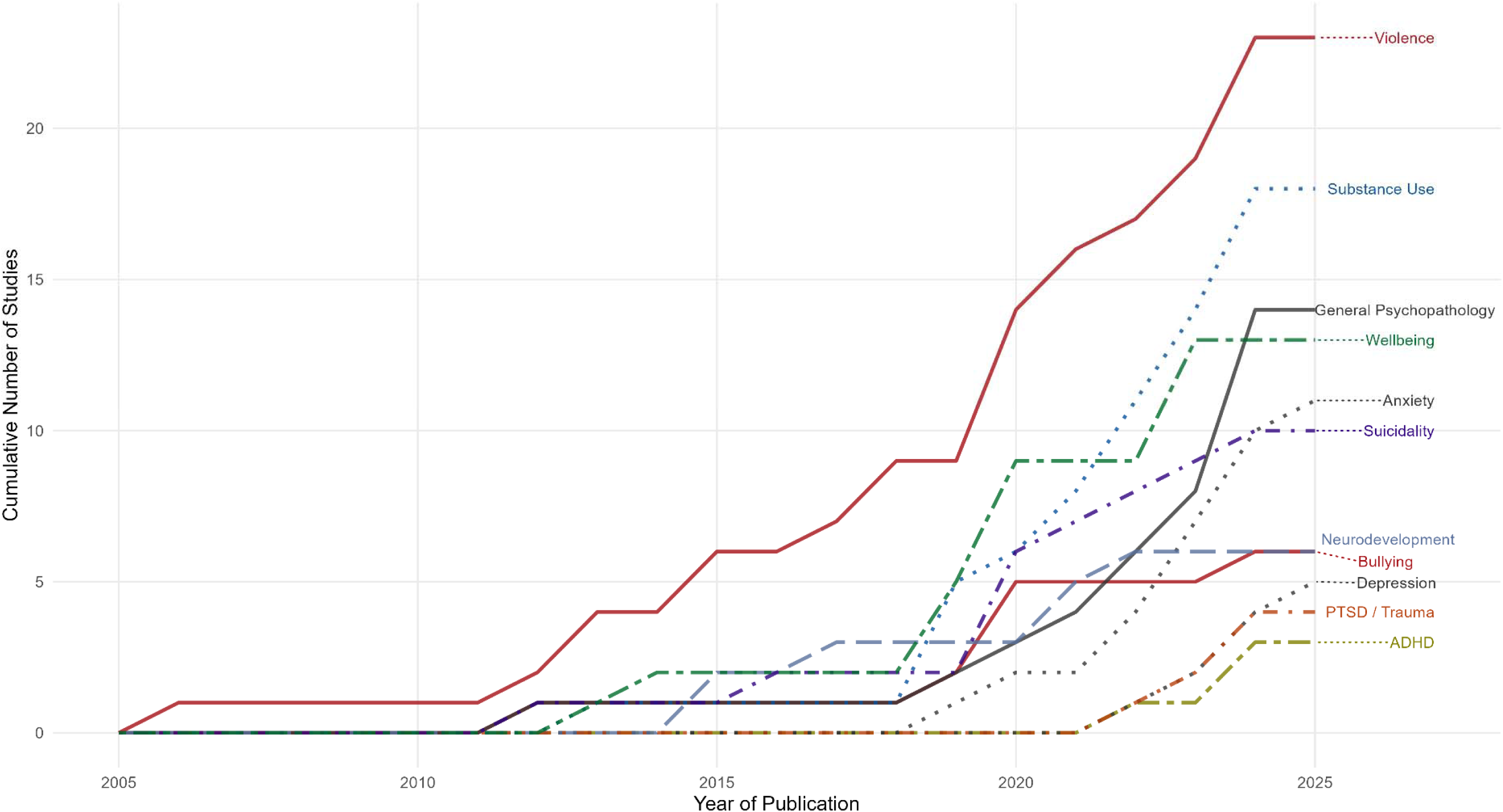
Cumulative trend of publications per diagnostic domain

International funding is also the primary source of support. There were several multinational surveys run by organizations such as the WHO,^26–47^ as well as national surveys sponsored by agencies like UNICEF^48^ and the NIMH.^49^ International development programs promoted 6 out of 7 intervention studies. Although more limited, national funding involves various public bodies. The National Demographic and Health Survey is a representative study which is implemented by the National Institute of Statistics, the Ministry of Health, and the National Institute for Health.^50^ Other contributing agencies include the National Council for Combating AIDS (CNCS)^21^ and the National Research Fund under the Ministry of Science, Technology, and Higher Education.^21^

### Prevalence

Table 1 presents studies based on structured clinical assessments (4) or health registers (8). There were no national level data on the prevalence of mental disorders, which relied on two local surveys employing structured clinical interviews and manualized diagnostic criteria. Among children and adolescents, a cross-sectional sample of 748 participants from Nampula estimated the prevalence of ADHD (13.4%). An assessment with 486 adolescents in Maputo reported rates for any anxiety disorders (17.5%), major depressive episode (8.5%), disruptive behavior disorder (6.6%), and ADHD (3.3%). Moreover, there was a non-probabilistic study with 110 individuals aged 1-20, reporting neurodevelopmental impairment according to clinical batteries (mild: 18.2%; moderate-to-severe: 0.9%). No clinical data was available on overall mental conditions nor estimating rates on ASD, eating disorders, and PTSD.

Health registers provided national data. A suicide rate of 0.65 per 100,000 people aged 5–19 was estimated based on a study collecting data from 2014 to 2019, with the highest rate of 1.53 among adolescents aged 15–19.^23^ All violent deaths are expected to be referred to forensic facilities, with three referral services operating in Mozambique at the time of the study. However, only absolute numbers were reported (70 suicides). Another study included all four reference hospitals in Mozambique, reporting cases of pediatric admissions due to injury caused by interpersonal violence, with absolute numbers of 5 occurrences in 2019 and 4 in 2020.

Table 2 outlines the remaining 46 studies estimating prevalence using screening instrument scores or responses to structured questionnaires. At national level, there were 6 probabilistic surveys with adolescents, with sample sizes ranging from 1,275 to 5,599 participants. These provided cut-off screening scores for two mental conditions (*generalized anxiety disorder: 6,3% - 18,51%; major depressive disorder: 6,6% - 10,25%*). It further informed several data points on substance use *(e.g: current tobacco use: 20.7%; current alcohol use: 13.7%)*, psychosocial symptoms *(e.g.: social isolation: 37.6%; suicidal ideation: 17.7%)*, socioeconomic vulnerabilities *(e.g.: out of school: 29%; orphanhood: 26,27%)*, and exposure to violence *(e.g: physical/emotional - last year: 24,86%; intimate partner violence suffered by girls - last year: 23,08%; lifetime sexual violence: 16,55%; frequent bullying - past month: 13,4%)*.

At regional level, three mental conditions were screened with a clinical sample of 2018 adolescents and young adults attending health services, estimating a higher burden of mental conditions among the 1096 HIV+ participants (*alcohol use disorder: 15.1%; PTSD: 14.7%; anxiety disorders: 10.3%; depression: 7.1%*). Probabilistic samples further included regional rates on low well-being scores *(31%, rural areas in Nampula, 1383 adolescents)* and mental symptom score bands *(e.g.: anxiety symptoms - high: 39,9%; depressive symptoms - severe: 14,2%; Gorongosa District, 794 children and adolescents).* There were no estimations for overall mental disorders, further lacking scores for ASD and eating disorders.

### Instruments

Table 3 shows cross-cultural adaptation and psychometric properties of 22 tools, including 2 diagnostic interviews, 12 screening scales, and 6 neurodevelopment tests. Supplementary Table 8 further lists 14 tools applied without formal adaptation or validation.

**Table 3.**
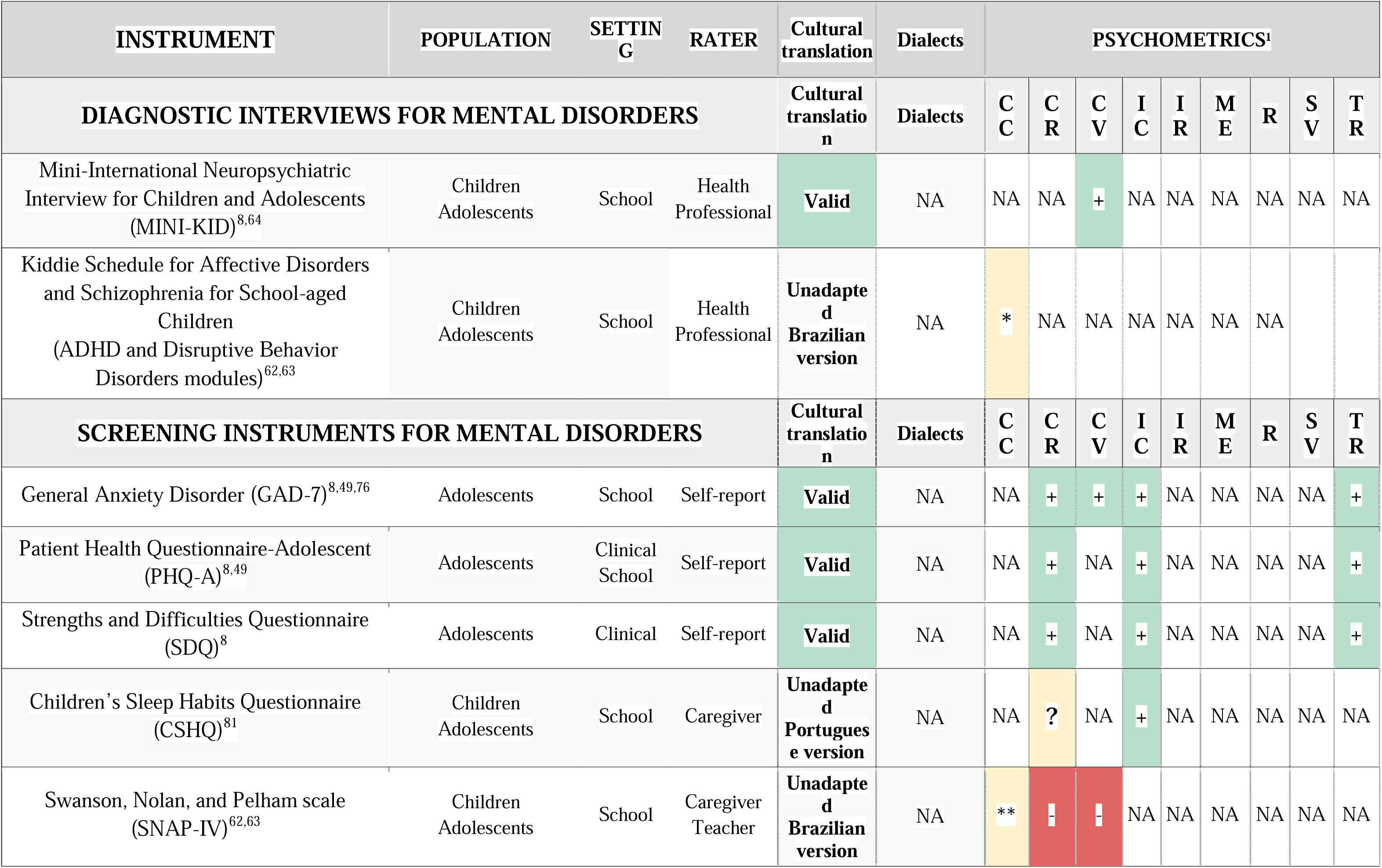
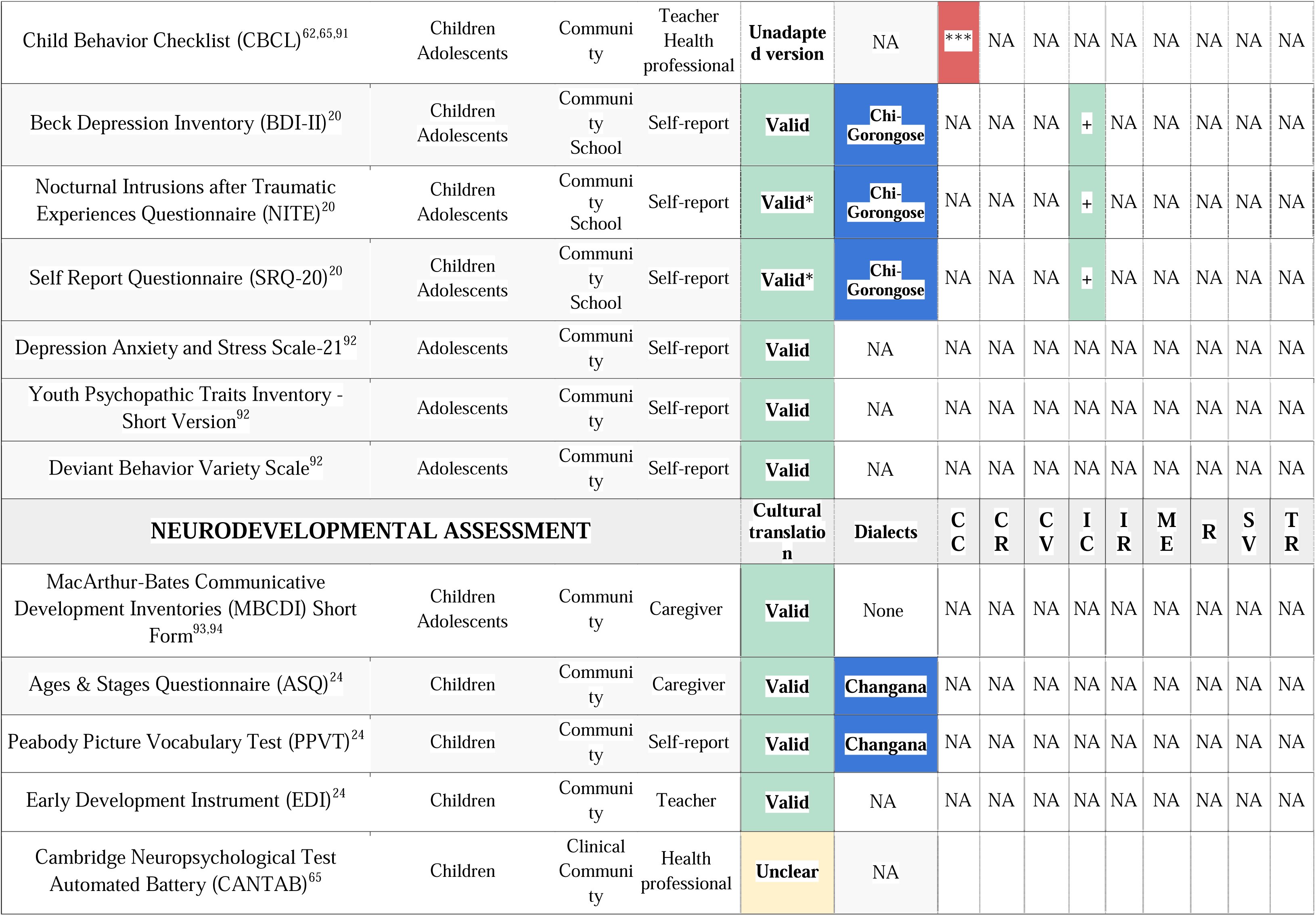
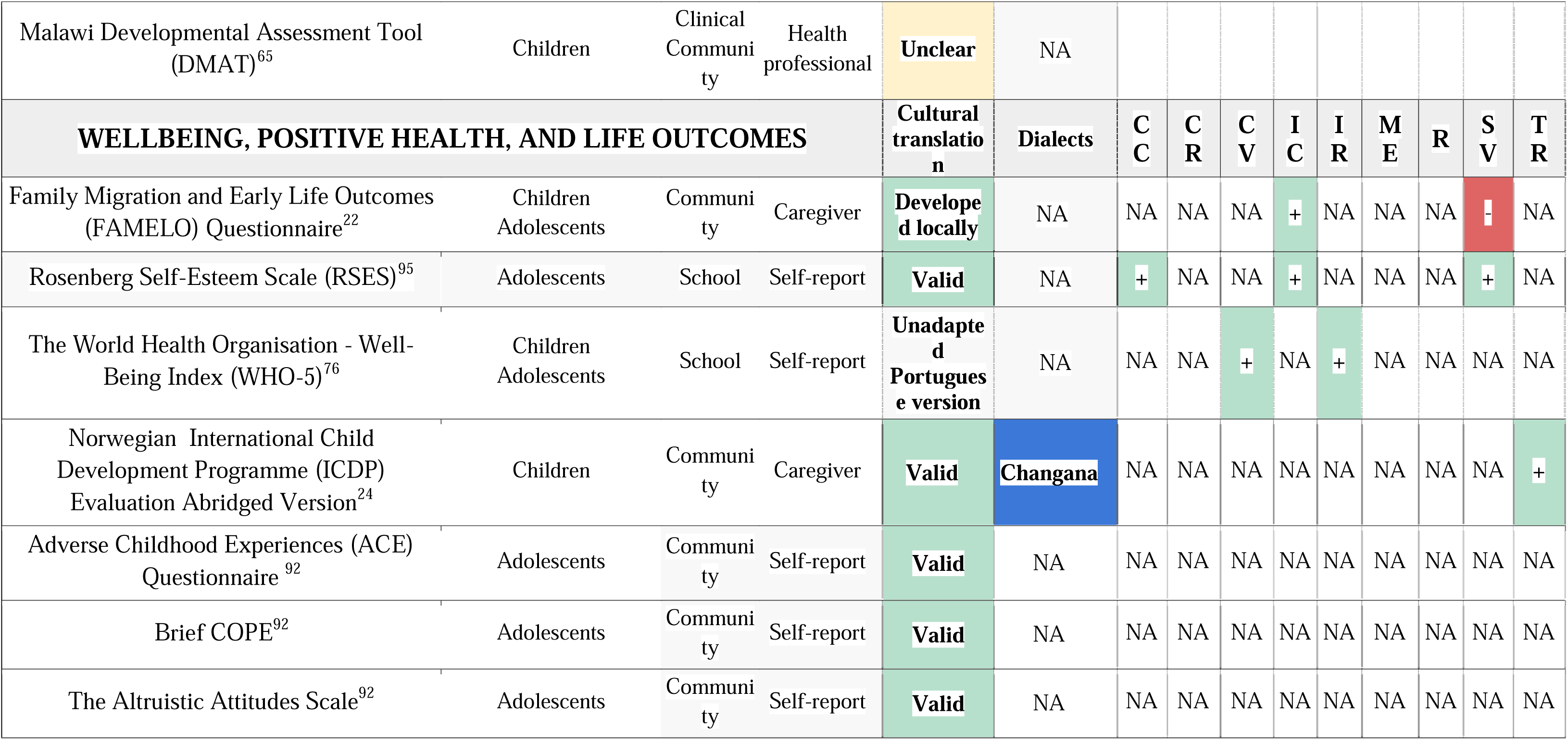
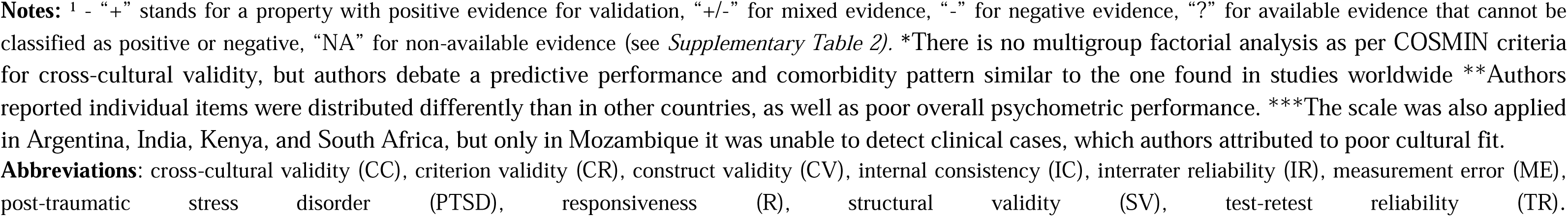
Instruments adapted or validated in Mozambique.

**Table 4.** Interventions.

| Intervention | Target | Design | Setting | Sample size<br>Age | Sample<br>description | Eff<br>ects | Results | RoB | Funding |
| --- | --- | --- | --- | --- | --- | --- | --- | --- | --- |
| Preschool program following a structured daily routine with play and learning activities<sup>24</sup> | Neurodevelo<br>pment | RCT | School | N = 1907<br>3 -5 years | Children from high-poverty rural areas | + | Significant gains in developmental skills (cognitive, communication, motor, and socio-emotional) at a cost of \$3 per child per month | High | World Bank Group - Education Global Practice Group |
| Adult–child communication (ACC) program developed by Go Girls! Initiative (GGI) with 11 sessions for caregivers<sup>96</sup> | Girl’s vulnerability to HIV | Controlled | Community | N = 603<br>(555 *caregivers*)<br>11 - 18 years | Adolescent girls in Zambezia and Nampula | ? | Girls reported improvement in relationship with parents, but hard outcomes were not assessed | High | President’s Emergency Plan for AIDS Relief (PEPFAR)<br>US Agency for International Development (USAID) |
| School personnel training with 4-day workshops to promote gender-equitable teaching and safer environments for girls<sup>97</sup> | Girl’s vulnerability to HIV | Controlled | School | N = 525<br>(67 *teachers*)<br>11 - 18 years | Teachers working with adolescent girls in Zambezia and Nampula | ? | Distal outcomes not assessed, no significant differences in students perception of teachers’ gender bias attitudes nor perceived safety at school | High | President’s Emergency Plan for AIDS Relief (PEPFAR)<br>US Agency for International Development (USAID) |
| Stop Violence Against Girls in School participatory program with multi-modal community activities<sup>25,85</sup> | Violence against girls | Uncontrolled | Community | N = 363<br>8 - 7 years | Girls in a rural area in Maputo | + | Increased percentage of girls reporting violence and showing gender-equatable attitudes<br>Increased the percentage of teachers with knowledge of laws, policies, and available support. | High | An ActionAid’s program supported by the UK’s Big Lottery Fund |
|  |  |  |  |  |  |  | No test of statistical significance. |  |  |
| International Child Development Programme (ICDP) with 10-12 weekly group sessions on parenting skills for caregivers<sup>98</sup> | Neurodevelopment | Controlled<sup>1</sup> | Community | N = 137<sup>2</sup><br>4.4 years<br>(SD 2.2) | Caregivers participating in the ICDP in Maputo | + | Statistically significant improvements on parents' self-efficacy, decrease on child conduct problems scores<br>Increased frequency of positive parenting practices |  | Part of the International Child Development Programme (ICDP), supported by multiple sources |
| Train community health workers to implement eight weekly psychodrama sessions<sup>99</sup> | Mental health promotion | Uncontrolled | Clinical | N = 35<br>15 - 24 years | Adolescents and young adults living with HIV attending supportive services in Beira | + | Statistically significant improvement in mental health knowledge and alexithymia | High | Part of Doctors with Africa CUAMM, supported by the Italian Agency for Development Cooperation (AICS) through the Global Fund project |
| Individual or pair activities for psychomotor and adaptive skills (twice-weekly 45-min sessions over 4 months)<sup>100</sup> | Neurodevelopment | Uncontrolled | Clinical | N = 6<br>6 - 13 years | Children with developmental disorders institutionalized in a rehabilitation center in Maputo | + | Significant improvement body awareness, gross motor skills, and adaptive skills | High | Not reported |

**Note:** <sup>1</sup>Post-hoc case-control design: 75 caregivers received the intervention, and 62 caregivers formed the control group. <sup>2</sup>The mean number of children per caregiver was 4.34, but analysis included only one child, focusing on the one closer to 4.4 years selecting
**Abbreviations:** RCT (Randomized Clinical Trial); RoB (Risk of Bias).

The Mini-International Neuropsychiatric Interview for Children and Adolescents (MINI-KID) was the only diagnostic interview that underwent cross-cultural adaptation, demonstrating positive evidence of construct validity. Among screening tools, three cross-culturally adapted instruments showed favorable psychometric performance across multiple properties *(Generalized Anxiety Disorder - GAD-7; Patient Health Questionnaire - Adolescent - PHQ-A; Strengths and Difficulties Questionnaire - SDQ)*. Other three tools were adapted to the local dialect *Chi-Gorongose*, yet psychometric assessment was restricted to internal consistency *(Beck Depression Inventory - BDI-II; Self Report Questionnaire - SRQ-20; Nocturnal Intrusions after Traumatic Experiences Questionnaire - NITE)*.

Five instruments were used in non-adapted versions originally developed for Brazil or Portugal, which was associated with poor psychometric performance: The Child Behavior Checklist (CBCL) failed to detect clinical problems in the Mozambican sample, despite performing adequately in other national arms of the same study conducted in Argentina, India, Kenya, and South Africa. The Swanson, Nolan, and Pelham scale (SNAP-IV) showed low predictive accuracy for ADHD diagnosis.

All neurodevelopment tests were cross-culturally adapted but lacked psychometric evaluation, with two tools translated to the local dialectic *Changana* (Ages & Stages Questionnaire - ASQ; Peabody Picture Vocabulary Test - PPVT). There was a lack of tools on ASD, eating disorders or self harm.

### Intervention studies

Table 4 presents seven intervention studies, including one RCT, three controlled trials, and three uncontrolled pre-post designs. Reports indicated a high risk of bias across all study designs. Programs consisted of 3 psychosocial interventions targeting neurodevelopment, 2 focusing on gender equality, and 1 on mental health promotion.

The RCT assessed a World Bank program among 1,907 preschool children in high-poverty areas. It demonstrated significant developmental gains with an implementation cost of approximately $3 per child per month. The single study focusing on mental health symptoms trained community health workers to deliver a psychodrama intervention for adolescents and young adults living with HIV+ Although lacking a control group, pre-post assessment with 35 participants showed statistically significant improvement in mental health knowledge and alexithymia.

## Discussion

This systematic review synthesizes evidence on child and adolescent mental health in Mozambique, encompassing 58 prevalence studies, 26 studies assessing 35 unique instruments, and 7 intervention trials. There are only regional data on diagnostic rates of mental disorders, with a survey with adolescents in Maputo (any anxiety disorder: 17.5%; major depressive episode: 8.5%; disruptive behavior disorder: 6.6%; ADHD: 3.3%) and another study with children and adolescents in Nampula (ADHD: 13.4%). Nationally representative surveys report mental health-related outcomes and risk factors, indicating that one in four adolescents experience physical or emotional violence annually, while 16.55% report lifetime sexual abuse, with particular concern regarding gender-based violence against girls. National suicide rates per age group could be estimated via epidemiological registers (15-19 years: 1.53; 10-14 years: 0.59; 5-9 years: 0.05). Only three assessment instruments (GAD-7, PHQ-A, and SDQ) have undergone cross-cultural adaptation and positive psychometric evaluation, while tools lacking cultural adaptation showed poor performance. Among interventions, only one RCT was identified, targeting neurodevelopment in high-poverty settings and showing positive outcomes and cost-effectiveness.

Data points from Mozambique indicate a higher prevalence of mental health disorders compared to findings from a similar Brazilian review^10^ and to global estimates (among adolescents: any anxiety disorder 4.34%, depressive disorders 2.9%, conduct disorders 2.63%, and ADHD 2%).^51^ Although suicide rates in Mozambique appear lower than then global averages (ages 15–19: 1.53; ages 10–14: 0.59 per 100,000), they are likely underestimated.^52^ The single national study was based on three referral services responsible for all violent death autopsies in the country.^23^ Moreover, the reliability of epidemiological and civil registration data in Mozambique is limited, with reports that nearly half of children under the age of five are not registered in official records.^3^ Similarly, other register-based studies in Mozambique are scarce and rely primarily on service-level data from isolated facilities, limiting their generalizability. An integrated national health information system that includes mental health outcomes is needed to strengthen epidemiological data and monitoring across the country.^53^

National-level data with adolescent samples indicate alarming rates of exposure to adversities and emotional and behavioral symptoms. In particular, the high prevalence of physical, emotional, and sexual violence stands out as a clear public health priority. These experiences are not only key social determinants of psychological distress and mental disorders,^54^ but also undermine healthy development and long-term prospects,^55,56^ potentially perpetuating cycles of violence and trauma across future generations.^57^ While current data do not represent children, it is likely that such exposures begin before adolescence. Surveys including infants, preschoolers, and school-aged children are recommended to support data-driven action from the earliest possible stages of development.^56^

We identified substantial gaps in the cross-cultural adaptation and psychometric validation of mental health instruments in Mozambique. While such gaps are common across non-English-speaking countries, they are particularly pronounced in Mozambique, where only 35 tools were identified (compared to 912 in Brazil and 261 in Greece). As a Portuguese-speaking country, Mozambique has often relied on instruments translated for use in Brazil or Portugal without further cultural adaptation, which was linked to poor psychometric performance. This highlights the need to adhere to culturally-sensitive practices in the country.

The volume and scope of scientific literature on child and adolescent mental health in Mozambique remain limited, presenting a recent research body in which most output emerged in the last decade. Reflecting the country’s socioeconomic challenges, existing studies largely focus on adversities such as maltreatment and violence, with some research clustered around neurodevelopment in vulnerable populations. While this underscores the elevated psychosocial burden and risk of mental disorders, few studies directly address mental health conditions. Prevalence data based on diagnostic criteria are restricted to a few disorders reported in only two localized samples in Maputo and Nampula, excluding other regions and rural areas. There are no studies on ASD, eating disorders, or self-harm, and no assessment tools are available for research or clinical practice on these conditions. Future efforts should prioritize closing the data gap on estimates of both overall and specific mental disorders among children and adolescents, improving representation of national and regional samples.

The majority of scientific literature has been produced through international collaborations and funded by development agencies. Few local experts appear as first or lead authors, and a single dissertation has been published by a Mozambican university. This reflects a scarcity of postgraduate programs and fragile academic infrastructure in the country. The need to strengthen national research capacity is acknowledged in the Strategy and Action Plan for Mental Health 2016–2026, which outlines an action front for developing research and human resources.^4^ The Department of Mental Health has established partnerships with institutions in Portugal, Brazil, and the United States to support postgraduate training in mental health.^58,59^ As a result, Mozambique now has 30 psychiatrists (10 PhDs, 6 Master’s), 693 psychologists (2 PhDs, 1 Master’s), and 41 occupational therapists (1 Master’s). Nevertheless, national universities remain weakly connected to medical specialization and offer few advanced training opportunities.^60^ Eduardo Mondlane University has a Master’s in Mental Health, and some doctoral programs in Biosciences have recently integrated mental health.^58^ Strengthening postgraduate training opportunities within the country is critical to building sustainable research capacity and a skilled mental health workforce.

This work builds on the methodological strengths of previous systematic reviews conducted in Greece and Brazil, employing a comprehensive search strategy and structured guidelines that provide best practices for appraising prevalence estimates, assessment instruments, and interventions. We further increased reach by including studies involving clinical samples and screening institutional reports and local catalogues. However, this review does not capture the totality of mental health research on children and adolescents in Mozambique. Additional data may be scattered across uncaptured sources, including conference abstracts,^61^ multinational assessments reporting only aggregate results, and non-academic sources. Given the limited scope of local research, these sources could represent a substantial body of evidence. Meta-analysis was not feasible due to the small number and significant heterogeneity of available studies.

The field of child and adolescent mental health is still emerging in Mozambique. Available data indicate a higher prevalence of mental disorders compared to global estimates, with public health priorities shaped by widespread socioeconomic adversity and alarming rates of gender-based violence. There are few available resources for screening mental health symptoms, and reliable assessment warrants cultural adaptation of instruments. Future research should target data gaps on prevalence rates of mental disorders across regional and national samples. There is an urgent need to strengthen local research capacity through investment in human resource development and the expansion of academic institutions.

## Ethical approvement

We obtained ethical approval from the Scientific Committee and Institutional Bioethics Committee for Health of the Faculty of Medicine at Eduardo Mondlane University and Maputo Central Hospital *(Comité Científico e Comité Institucional de Bioética para Saúde da Faculdade de Medicina da Universidade Eduardo Mondlane e Hospital Central de Maputo*) [Reference: CIBSFM&HCM/42/2025]. Although this study did not involve direct participation of human subjects, national regulations in Mozambique require local ethical approval for all review studies.

## Data Availability

All data produced in the present work are contained in the manuscript

