## Supplementary for "The science of child and adolescent mental health in Mozambique: a nationwide systematic review"


Supplementary Table 1 - Search strategy and query per database 2
Supplementary Table 2 - Prevalence studies: extracted information for each estimate 4
Supplementary Table 3 - Data extracted for each instrument reported at each study 5
Supplementary Table 4 - Instruments: evaluation of psychometric properties and language 7
Supplementary Table 5 - Synthesis of each instrument property: coding criteria 8
Supplementary Table 6 - Intervention studies: extracted information 9
Supplementary Table 7 - Reasons for exclusion 10
Supplementary Table 8 - Instruments that were applied in Mozambique (without adaptation or validation) 11

Supplementary Table 1 - Search strategy and query per database

| **Database / Repositoire** | **Strategy** | Query |
|---|---|---|
| Pubmed<br>PsycInfo<br>Web of Science<br>CINAHL | Full query English terms (syntax adapted per databases)<br><br>Results are exported and deduplicated<br><br>Two independent reviewers perform primary (title/abstract) and secondary screening (full text). | *(adolesc* OR preadolesc* OR pre-adolesc* OR child* OR boy OR girl OR infant* OR juvenil* OR minors OR paediatri* OR pediatri* OR pubescen* OR puberty OR school* OR student* OR teen* OR young OR youth* OR class* OR orphan* OR high-school OR "high school" OR preschool* OR pre-school*)*<br>***AND*** *(Mozambique OR Mozambican OR Moçambique)*<br>***AND*** *(Mental OR "Mental health" OR "Mental Disorder" OR Psychiatr* OR Psycho* OR behavior OR Mood OR Emotion OR Cognitive OR Neurocognitive OR Autism OR Enuresis OR Encopresis OR ADHD OR "Attention Deficit Hyperactivity Disorder" OR Intellectual disability OR development delay OR neurodevelopment OR Oppositional Defiant OR Conduct OR Depression OR depressive OR Bipolar OR Suicide OR Suicidality OR Selfharm OR Obsessive-compulsive OR Trauma OR PTSD OR Mutism OR "Substance use" OR Cannabis OR Alcohol OR Tobacco OR Anorexia OR Bulimia OR "Eating disorder" OR "Personality Disorder" OR Schizophrenia OR Psychosis OR "Quality of life" OR wellbeing OR "learning disorder" OR "learning delay" OR anxi* OR phob* OR panic)** |
| African Index Medicus | Simplified query in English and Portuguese<br><br>Single-reviewer screening of all results | *Mozambique OR Moçambique* |
| Google Scholar | Simplified queries in English and Portuguese<br><br>A single reviewer screening results until reaching 100 sequential articles without novel inclusions | ***1. English search:*** *Mozambique (Mental OR Psychiatry OR Psychology) (Child OR Adolescent OR school)*<br><br>***2. Portuguese search****: Moçambique (Mental OR Psico* OR Psiq*) (Criança OR adolescente OR escola OR infancia OR juventude)* |

| | | |
|---|---|---|
| Universidade Eduardo Modlane (UEM) | Simplified query in Portuguese<br><br>Single-reviewer screening of all results | *adolescente OR pré-adolescente OR criança OR menino OR menina OR bebê OR juvenil OR menores OR pediátrico OR puberdade OR escola OR estudante OR adolescente OR jovem OR juventude OR órfão OR "ensino médio" OR "escola secundária"* |
| Instituto Nacional de Saúde (INS)<br>Universidade Lúrio (UniLurio)<br>Revista Médica de Moçambique<br>Revista Moçambicana de Ciência de Saúde | Single-reviewer screening of all publications | |

Supplementary Table 2 - Prevalence studies: extracted information for each estimate

| Topic | Variable |
|---|---|
| **Study details** | First author |
| **Study details** | Year of publication |
| **Study details** | Study description |
| **Study details** | Region |
| **Data of each estimate** | Diagnostic domain |
| **Data of each estimate** | Condition or construct |
| **Data of each estimate** | Year of data collection |
| **Data of each estimate** | Sampling/representativeness |
| **Screening sample** *(if available)* | Instrument |
| **Screening sample** *(if available)* | Cut-off for positive |
| **Screening sample** *(if available)* | Informant |
| **Screening sample** *(if available)* | Recruitment method |
| **Screening sample** *(if available)* | Sample size |
| **Screening sample** *(if available)* | Response rate |
| **Screening sample** *(if available)* | Age range |
| **Screening sample** *(if available)* | Percentage of males |
| **Screening sample** *(if available)* | Number positive |
| **Diagnostic sample** | Sample description |
| **Diagnostic sample** | Sample size |
| **Diagnostic sample** | Response rate |
| **Diagnostic sample** | Percentage of males |
| **Diagnostic sample** | Assessment instrument |
| **Diagnostic sample** | If instrument includes interview |
| **Diagnostic sample** | Informants |
| **Diagnostic sample** | Diagnostic criteria |
| **Diagnostic sample** | If diagnosis requires functional impairment |
| **Diagnostic sample** | Definition of functional impairment |
| **Diagnostic sample** | Type of prevalence |
| **Results** | Prevalence estimate (SD or 95% CI) |

#### Supplementary Table 3 - Data extracted for each instrument reported at each study

| **Topics / Procedures** | **Extracted information** |
|---|---|
| General information | Details on the study, instrument, and sample |
| Study procedures | Whether it develops, validates and/or translates, or only applies an instruments |
| Development quality* | Methodological quality** |
| Content validity quality* | Methodological quality** |
| Instrument translation | The presence of procedures such as back-and-forth translation, independent translators, expert committee assessment, and pilot testing |
| Structural validity | Type of analysis (e.g., exploratory factor analysis, principal component analysis, or confirmatory factor analysis)<br>How many factors best accounted for how much of the variance<br>Results of statistical procedures (e.g., Root Mean Square Error of Approximation (RMSEA), Comparative Fit Index (CFI), Standardized Root Mean Residuals (SRMS))<br>Sample size and methodological quality** |
| Internal consistency | Cronbach's alpha or equivalent measures<br>Sample size and methodological quality** |
| Cross-cultural validity | Multi-group confirmatory factor analysis with measurement invariance evaluation<br>Sample size and methodological quality** |
| Inter-rater reliability | Intraclass correlation coefficient, pearson or spearman correlation, kappa score, or equivalent statistics<br>Sample size and methodological quality** |
| Test-retest reliability | Intraclass correlation coefficient, pearson or spearman correlation, kappa score, or equivalent statistics<br>Sample size and methodological quality** |
| Measurement error | Statistics on the patient's score error (e.g., standard error measurement, smallest detectable change, or limits of agreement)<br>Sample size and methodological quality** |
| Criterion validity | Performance of the instrument against a gold standard (e.g., area under the curve, sensitivity, and specificity)<br>Sample size and methodological quality** |
| Construct validity | Correlation coefficients with other instruments<br>Sample size and methodological quality** |

| Topics / Procedures | Extracted information |
|---|---|
| Responsiveness | Detection of statistically significant differences in measures over time (e.g., after an intervention)<br>Sample size and methodological quality** |

**Notes:** This table is reproduced from Marchionatti et al. (2024).[10] *Only for studies reporting instrument development. **Classified as very good, adequate, doubtful, inadequate, or not applicable, following the Consensus-based Standards for the Selection of Health Measurement Instruments (COSMIN)[16] guidelines

#### Supplementary Table 4 - Instruments: evaluation of psychometric properties and language

| Code | Criteria for each psychometric property evaluation |
|---|---|
| + | The property was measured and the study provides positive evidence according to standard definitions from COSMIN, except for adapted definition for responsiveness<br>• **Structural validity** with CFA with CFI >0.95/RMSEA <0.06<br>• **Internal consistency** with Cronbach's alpha > 0.7<br>• **Reliability** with Pearson's *r*, ICC or weighted Kappa ≥ 0.70<br>• **Cross-cultural validity** with no differences between group factors; reliability with ICC or Kappa >= 0.70<br>• **Measurement error** with SDC or LoA < MIC<br>• **Criterion validity** with correlation with gold standard >= 0.70<br>• **Construct validity** with correlations superior 0.10 or 0.5, depending on the hypothesis<br>• **Responsiveness** statistically significant differences between time-points |
| - | The property was measured and the study provides negative evidence according to standard definitions (e.g., Cronbach's alpha < 0.7 for internal consistency, confirmatory factor analyses with CFI <0.95/RMSEA >0.06 for structural validity) |
| +/- | Mixed outcomes that may be classified either as "+" or "-" |
| ? | Information was provided for the property, but it is insufficient to ascertain it in previous categories |
| NA | Information was not available for that property |
| **Code** | **Criteria for study language and procedures** |
| **D** | Development study (in Portuguese or other local language) |
| + | Study translates the instrument with structured procedures as back-and-forth translation, independent translators, expert committee assessment, and pilot testing |
| - | Study translated the instrument without structured procedures |
| ? | Employs a previously translated version of the instrument |

**Notes**: This table is reproduced from Marchionatti et al. (2024).[10]. The summary appraisal table evaluates data from the extraction table, with each entry corresponding for each instrument procedure at each study. Refer to Consensus-based Standards for the selection of health Measurement Instruments Manual (COSMIN) for details of criteria.[16] For responsiveness, we considered sufficient ("+") evidence of validation if the instrument presents statistically significant differences in a clinical trial. This procedure was chosen because studies did not provide the area under the curve in this property, which was the original parameter in the COSMIN manual. **Abbreviations**: Confirmatory Factor Analysis (CFA), Comparative Fit Index (CFI), Intraclass Correlation Coefficient (ICC), Limits of Agreement (LoA), Minimal Important Change (MIC), Root Mean Square Error of Approximation (RMSEA), Smallest Detectable Change (SDC).

Supplementary Table 5 - Synthesis of each instrument property: coding criteria

| Code | Criteria for psychometric properties |
|---|---|
| + | Available reports with positive validation results on that psychometric property |
| +/- | Available reports with both positive and negative validation results on that psychometric property |
| - | Available reports with negative validation results on that psychometric propertie |
| ? | Available reports for that psychometric property that cannot be classified as positive or negative |
| NA | No available reports for that psychometric property |
| **Code** | **Criteria for study language and procedures** |
| D | An instrument developed in Portuguese or other local language with reports on its development |
| PT-BR | Instruments originally in Portuguese or other local language without reports on its development |
| P | Instruments with a previous translation to Portuguese or other local language, without reports on its procedures |
| + | Instruments that were translated or cross-culturally adapted to Portuguese or other local language with at least one of the following procedures: back-and-forth translation, independent translators, expert committee assessment, and pilot testing |
| - | Instruments that were freely translated to Portuguese or other local language |

**Note:** This table is reproduced from Marchionatti et al. (2024).[10] Synthesis table aggregates all information reported for each instrument across extracted studies.

Supplementary Table 6 - Intervention studies: extracted information

| Field | Details |
|---|---|
| Year | |
| Author | |
| Condition | |
| Design | RCT, controlled, uncontrolled, adaptation or development protocol |
| Type of intervention | Psychosocial, life style or biomedical |
| Experimental interventions | |
| Control Interventions | |
| Target construct | |
| Primary outcome | |
| Primary outcome measurement | |
| Secondary outcome | |
| Secondary outcome measurement | |
| Other measures | |
| Population description | |
| Sample (age, gender, race, location) | |
| Inclusion criteria | |
| Exclusion criteria | |
| Method of recruitment | |
| Allocation methods | Include unit of allocation (cluster or individual) |
| Size per group | Intervention and control, total number randomized |
| Baseline imbalances | |
| Duration of intervention | |
| Withdrawals and exclusion | |
| Other treatment received | |
| Time points measured | |
| Informant | |
| Effect size | Controlled and uncontrolled |
| Results | |
| Demonstrated efficacy? | |
| Risk of bias[18,19] | The revised Cochrane tool for randomized trials (RoB 2)<br>The Joanna Briggs Institute (JBI) tool for non-randomized designs |
| Funding | |

### Supplementary Table 7 - Reasons for exclusion

| **Primary Screening** |
|---|
| Wrong outcome: 3406 |
| Wrong population: 157 |
| Wrong study design: 33 |
| Wrong publication type: 15 |
| Background article: 1 |
| **Secondary Screening: Prevalence Studies** |
| Wrong population: 206 |
| Wrong outcome: 65 |
| Wrong study design: 31 |
| Wrong publication type: 12 |
| Background article: 12 |
| Full report unavailable 7 |
| **Secondary Screening: Instrument Studies** |
| Wrong population: 132 |
| Wrong study design: 124 |
| Only applies an instrument already Included: 63 |
| Wrong outcome: 32 |
| Background article: 8 |
| Wrong publication type: 5 |
| **Secondary Screening: Intervention Studies** |
| Wrong study design: 28 |
| Wrong population: 13 |
| Wrong publication type: 5 |
| Wrong outcome: 4 |
| Full report undeniable: 3 |
| Background article: 1 |

Supplementary Table 8 - Instruments that were applied in Mozambique (without adaptation or validation)

| Tools applied without providing data on psychometric validation | |
|---|---|
| **Assessment instruments** | **Questionnaires from global surveys** |
| Children's Global Assessment Scale (CGAS)[62,63] | Demographic and Health Surveys (DHS)[101,102] |
| Alcohol, Smoking, and Substance Involvement Screening Test (ASSIST)[62] | Global Youth Tobacco Survey (GYTS)[103] |
| Pediatric Quality of LIfe Inventory (PedsQL)[104] | WHO Multi-country Survey of Women's Health and Domestic Violence against Women[84] |
| Eysenck Personality Questionnaire - Revised (EPQR)[88] | Global School Health Survey (GSHS)[105] |
| Beck Anxiety Inventory (BAI)[88] | Determined, Resilient, Empowered, AIDS-Free, Mentored, and Safe (DREAMS) Questionnaire[77] |
| Bateria Psicomotora (BPM)[100] | |
| Portuguese Adaptive Behavior Scale (PABS)[100] | |
| Child PTSD Symptom Scale for DMS-V (CPSS-V )[49] | |
| Conflict and Tactics Scale (CTS)*[90] | |

**Note:** Chaquisse 2018 Sexual and physical: Conflict and Tactics Scale (CTS) previously adapted to Mozambique, but for adult populatio
